# High dose-rate brachytherapy of localized prostate cancer converts tumors from cold to hot

**DOI:** 10.1101/2020.03.02.20030346

**Authors:** Simon P Keam, Heloise Halse, Thu Nguyen, Minyu Wang, Nicolas Van Kooten Losio, Catherine Mitchell, Franco Caramia, David J Byrne, Sue Haupt, Georgina Ryland, Phillip K Darcy, Shahneen Sandhu, Piers Blombery, Ygal Haupt, Scott G Williams, Paul J Neeson

**Author notes:** **Corresponding author:** Paul J Neeson, Cancer Immunology Research, Peter MacCallum Cancer Center, 305 Grattan St, Parkville, Melbourne, 3000, Australia. Denotes shared first author (Simon P Keam, Heloise Halse). Denotes shared senior author (Scott G Williams, Paul J Neeson). **Conflict of Interest Statement:** The authors have no conflicts of interest to declare. **Author contributions** S.P.K., H.H., S.G.W. and P.J.N conceived and designed the study. S.P.K., H.H., T.N., N.V.K.L., D.J.B. and G.R. performed experiments. S.P.K., M.W. and F.C. performed bioinformatic and statistical analyses. C.M. performed uropathological inspection. S.P.K. and P.J.N. drafted the manuscript. S.P.K., H.H., P.J.N., SH., Y.H., C.M., D.B., G.R., P.K.D., S.S. and P.B. critically revised the manuscript. **Funding** This study was funded by the PeterMac Foundation, the Victorian Cancer Agency, a Prostate Cancer Foundation USA (Creativity award), Cancer Council Victoria (Grant-In-Aid) and an NHMRC Program Grant. **Availability of data and materials** The datasets used and/or analysed during the current study are available from the corresponding author on reasonable request.

## Abstract

Prostate cancer (PCa) has a profoundly immunosuppressive microenvironment, we hypothesized that radiation therapy would break this immune suppression. To investigate this, we performed high-throughput immune cell subset analysis, and gene expression profiling of pre-versus post-radiation tissue in a cohort of patients with localized disease that received high dose-rate brachytherapy (HDRBT). We resolved tumor and non-tumor zones in our spatial analysis to explore what drives the immunological response. Nanostring immune profiling revealed numerous immune checkpoint molecules were stimulated in response to HDRBT (e.g. B7-H3, CTLA4, PDL1 and PDL2). A published 16-gene tumor inflammation signature (TIS) gene profiling of immune activation states (high:*hot*, intermediate and low:*cold*) showed that most tissues possessed a low TIS pre-HDRBT. Crucially, HDRBT converted 80% of these ‘cold’-phenotype tumors into an ‘intermediate’ or ‘hot’ class. We used digital spatial profiling to show these HDRBT-induced changes in prostate TIS scores were derived from the non-tumor regions. Furthermore, these changes in TIS were also associated with pervasive changes in immune cell density and spatial relationships – in particularly between T cell subsets and antigen presenting cells. We identified increased density of CD4^+^ FOXP3^+^ T cells, CD68^+^ macrophages and CD68^+^ CD11c^+^ dendritic cells in response to HDRBT. The only subset change in tumor zones was PDL1^+^ macrophages. While these immune responses were heterogeneous, HDRBT induced significant changes in immune cell associations, including a gained T cell and HMWCK^+^ PDL1^+^ interaction in tumor zones. In conclusion, we showed HDRBT has a clear impact in converting “cold” prostate tumors into more immunologically activated “hot” tissues, with accompanying spatially-organized immune infiltrates and signaling changes. Understanding and potentially harnessing these changes will have widespread implications for the future treatment of localized PCa and the possible use of combination immunotherapies.

## Introduction

Standard curative-intent treatment options for localized prostate cancer include radical prostatectomy or radiotherapy [1]. Radiation therapy is delivered using external beam radiotherapy (EBRT) or via radioactive sources implanted within the prostate (brachytherapy). High-dose-rate brachytherapy (HDRBT) uses a temporary prostate implant to deliver high doses of radiation over a short time, achieving a high tumor dose while minimizing radiation exposure to nearby normal tissue, while often employing an additional short course of EBRT to treat surrounding tissues.

HDRBT enables substantially higher doses (upwards of 10Gy per fraction usually) to be delivered to the prostate than those typically used with EBRT (1.8-2.0Gy per fraction) and exploits the known sensitivity of PCa to such large fractional doses [2]. This relative resistance to conventional low doses of EBRT has historically been attributed to an enhanced ability of PCa to repair sublethal DNA damage [3]. This results in excellent responses for the majority of patients, however at least a third of men will ultimately relapse [4] and progress to advanced and incurable disease.

Normal prostate has an immune infiltrate characterized by intra-epithelial lymphocytes in the prostate glands, and low numbers of lymphocytes and macrophages distributed throughout the interstitial tissue [5]. Localized PCa has an immunosuppressed phenotype, where tumor-associated lymphocytes are present at the tumor margin, with only sparse numbers within the tumor [6]. This suggests that an anti-tumor immune response exists in these patients, but is held in check by the immunosuppressed prostate tumor.

In addition to direct effects on tumor cells and supporting stromal cells, radiotherapy can provoke immunological effects [7]. Clinical evidence of this can be found in case studies where distant untreated metastases undergo apparent spontaneous regression post-radiotherapy of an index lesion; an abscopal effect [8]. This is thought to be mediated via a systemic immune response triggered by the initial RT. The mechanisms implicated include immunogenic tumor cell death, antigen cross presentation and T cell priming to tumor-derived antigens [9]. However, there is pre-clinical evidence that hypo- and hyper-fractionated RT have different immunological effects on tumor tissue. Specifically, that high dose-rate hypo-fractionated radiation, which includes HDRBT, can break immunosuppressive tumor microenvironments, promote immune cell infiltration, and induce anti-tumor responses [10]. However, establishing these differences in clinical samples has been difficult due to heterogeneity in dose regimens and RT delivery methods.

Presently, it is also unclear whether the clinical response seen with HDRBT results from direct action toward PCa cells, cancer associated fibroblasts, the immune system or any combinations of these. For this reason, we have specifically explored these three potential response mechanisms to HDRBT in a cohort of patients with localized PCa. We previously revealed PCa stromal cell gene expression and proteomic responses to HDRBT as well as dosimetric features [11-13]. In this current study, we used the same patient cohort to evaluate localized PCa immune response to HDRBT. To do this, we assessed a cohort of 24 paired pre- and post-HDRBT PCa samples for changes in tumor inflammation gene signatures and multiplex IHC. Contrasting this information with alterations in immune cell densities, cell associations and spatial relationships revealed the capacity for HDRBT to fundamentally alter the tumor immune microenvironment.

## Materials and Methods

Complete methods and materials, including supporting quality control information are provided in Supplementary Information.

### Patient cohort and sample collection

**Table 1** shows the clinical characteristics of 24 patients with localized PCa analyzed in this study. Patients had two HDRBT treatments performed 14 days apart with a 10Gy fraction prescribed to the target volume (whole prostate) on each occasion. Full details of radiation therapy delivery, including sample processing, histopathological assessment, and ethics approval are provided in Supplementary Information. All participants provided consent covering tissue research as part of a prospective tissue collection study for prostate radiobiology research, approved by the Human Research Ethics Committee at the Peter MacCallum Cancer Centre (PMCC; HREC approvals 10/68, 13/167, 18/204).

**Table 1:** Clinical characteristics of cohort.

| Gleason Grade Group | T-stage | Number of patients | Age (mean ± SD) | PSA (ng/ml) | Neoadjuvant ADT | Relapse |
| --- | --- | --- | --- | --- | --- | --- |
| 2 | 1c | 4 | 59.3 ± 7.9 | 6.1 ± 1.6 | | |
| | 2a | 2 | 70.5 ± 3.5 | 7.7 ± 4.6 | | |
| | 2b | 2 | 70.0 ± 1.4 | 8.8 ± 1.9 | | |
| | Total | 8 | 64.8 ± 8.0 | 7.1 ± 2.5 | | |
| 3 | 1c | 5 | 63.0 ± 4.5 | 21.6 ± 32.8 | Yes | Yes |
| | 2b | 3 | 65.3 ± 4.2 | 12.1 ± 6.8 | | |
|  | 2c | 1 | 70.0 | 11.3 | Yes |  |
|  | 3a | 1 | 67.0 | 11.2 |  | Yes |
|  | 3b | 1 | 57.0 | 80.0 | Yes |  |
| | Total | 11 | 64.1 ± 4.7 | 22.4 ± 28.8 | 3 | |
| 4 | 1c | 1 | 76.0 | 27.7 | Yes |  |
|  | 2a | 1 | 70.0 | 6.1 |  |  |
| | Total | 2 | 73.0 ± 4.2 | 16.9 ± 15.3 | 1 | |
| 5 | 2a | 1 | 66.0 | 6.4 |  |  |
|  | 2b | 1 | 73.0 | 7.2 | Yes |  |
|  | 3b | 1 | 67.0 | 10.3 | Yes | Yes |
| | Total | 3 | 68.7 ± 3.8 | 8.0 ± 2.1 | 2 | |
| | | 24 | 65.6 ± 6.2 | 15.1 ± 20.7 | 6 | 3 |

### Multiplex IHC

3 µm FFPE tissue sections from each biopsy were mounted on an adhesive slide, deparaffinized and rehydrated by serial passage through changes of xylene and graded ethanol for multiplex immunohistochemistry staining with the T cell Panel and Pan Immune Panel as described previously [14]. Full details for imaging, cell segmentation, data processing and quality control are provided in Supplementary Information. An example of the experimental pipeline is shown in **Supplementary Figure S1** and quality control data are shown in **Supplementary Figures S2, S3** and **S4**. Further information is provided in the Supplementary material.

### Nanostring and 3’ RNAseq immune gene expression profiling

RNA was isolated and purified from 10 µm tissue sections using the RNeasy FFPE kit (Qiagen) according to the manufacturer’s instructions. Full details for nCounter PanCancer Immune Profiling Panel and 3’ RNAseq analyses are provided in the Supplementary Information.

### Digital Spatial Profiling

3 µm serial sections of post-HDRBT biopsies from patients RB019 and RB023 were processed by Nanostring Technologies using the ImmunoOncology (IO) RNA and protein panels and analyzed using a GeoMx. Sections were stained prior to ROI selection (12 ROIs per section) with CD3, CD68, Pan-Cytokeratin (PanCK) and DAPI to enable rational selection of regions. Raw data was normalized to both External RNA Control Consortium (ERCC) spike-in and signal-to-noise control. These normalized data were used for generation of TIS signature (mean of all normalized values in TIS signature).

### Data Analysis, bioinformatics and statistical considerations

Full details for data analyses, as well as statistical tests and power calculations are provided in the Supplementary Information.

## Results

Clinical and histopathological features of the twenty-four patients who underwent HDRBT for localized prostate cancer are summarized in **Table 1**. Forty eight tissue samples were obtained from these patients, and analyzed as depicted in **Figure 1A**. H&E sections from these paired biopsies were viewed by an experienced uropathologist to mark the location of tumor-bearing tissue within the core (**Figure 1B**). Immune gene expression changes in response to radiation were evaluated using the Nanostring PanCancer Immune Profiling panel, digital spatial profiling (DSP) and the TIS gene signature. Serial sections were used as input for multiplex IHC (mIHC) on two different multiplex panels (pan–immune or T cell-specific) to enumerate 12 distinct cell subsets (refer to **Table 2** for cell phenotypes). Using the multiplex IHC data, we derived immune subset density (cells per mm^2^) and spatial characteristics (median cell-to-cell distance), and correlated this with immune gene expression data, immune network signaling and the Nanostring tumor inflammation score.

**Table 2:** Immune subsets assessed by multiplex IHC.

| Marker expression | Panel | Cell subset | Abbreviation |
| --- | --- | --- | --- |
| **CD3<sup>+</sup>CD4<sup>+</sup>CD8<sup>-</sup>** | T cell | T-helper | T-helper |
| **CD3<sup>+</sup>CD8<sup>+</sup>CD4<sup>-</sup>** | T cell | CD8<sup>+</sup> T cells | CD8<sup>+</sup> T cells |
| **CD3<sup>+</sup>CD4<sup>+</sup>FOXP3<sup>+</sup>** | T cell | Regulatory T cell | Treg |
| **CD3<sup>+</sup>CD4<sup>-</sup>CD8<sup>-</sup>** | T cell | Double negative T cell | DNT |
| **CD3<sup>+</sup>** | Pan Immune | Pan T cell | T cell |
| **CD20<sup>+</sup>** | Pan Immune | B cell | B cell |
| **CD68<sup>+</sup>** | Pan Immune | Macrophage | φ |
| **CD68<sup>+</sup>CD11c<sup>+</sup>** | Pan Immune | Dendritic cell | DC |
| **CD68<sup>+</sup>PDL1<sup>+</sup>** | Pan Immune | PDL1<sup>+</sup> Macrophage | φ<sup>PDL1+</sup> |
| **CD68<sup>+</sup>CD11c<sup>+</sup>PDL1<sup>+</sup>** | Pan Immune | PDL1<sup>+</sup> Dendritic cell | DC<sup>PDL1+</sup> |
| **HMWCK<sup>+</sup>PDL1<sup>+</sup>** | Pan Immune | PDL1<sup>+</sup> Basal cells | HMWCK<sup>+</sup>PDL1<sup>+</sup> |
| **HMWCK<sup>-</sup>PDL1<sup>+</sup>** | Pan Immune | PDL1<sup>+</sup> Non-basal cells | HMWCK<sup>-</sup>PDL1<sup>+</sup> |

**Figure 1:**
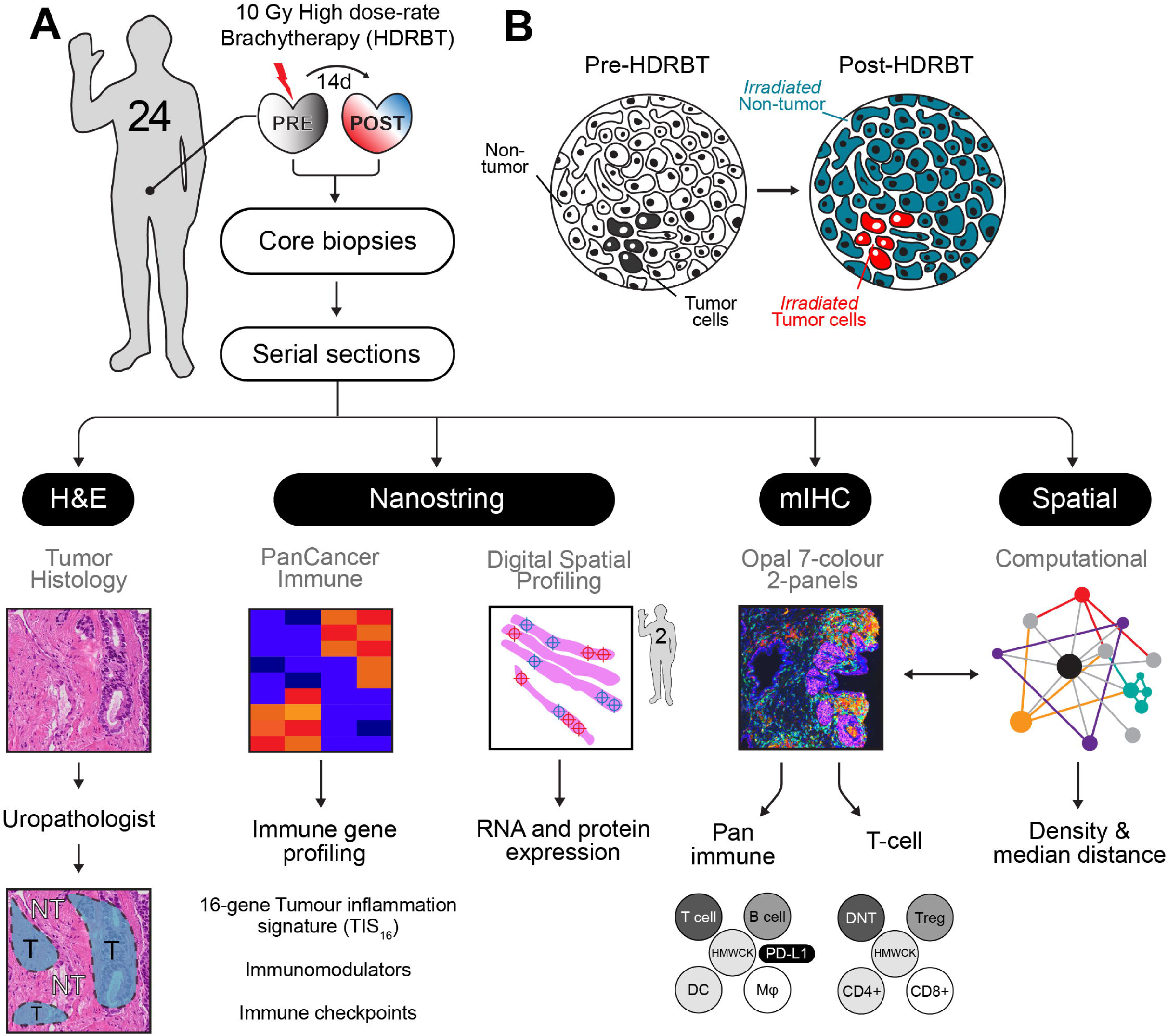
**Overview of experimental plan**. (A) Twenty four patients with localised prostate adenocarcinoma were treated with a single 10 Gy dose of high-dose-rate brachytherapy (HDRBT) with image guided biopsies taken immediately prior to and two weeks post-HDRBT. Following formalin fixation and sectioning, samples were processed for (i) histopathological examination and tumor zone categorisation, (ii) Nanostring nCounter Pan-cancer immune gene panel RNA profiling, (iii) digital spatial profiling, (iv) multiplex IHC, and (v) computation spatial analysis. (B) Schematic showing tissue zones categorised by histopathology and incorporated into subsequent immune density analyses. T: tumor, NT: non-tumor, HDRBT: high dose-rate brachytherapy, mIHC: multiplex immunohistochemistry, HMWCK: high molecular weight cytokeratin, DC: dendritic cells, MΦ: macrophage, Treg: regulatory T cells.

### Profiling the transcriptional activation response of PCa to HDRBT

We hypothesized that HDRBT alters the local immune signaling network of PCa, and that this would be evident in changes in immune activation and signaling at the transcriptional level. We therefore used an established immune profiling gene expression platform and robust signature of clinically-relevant immune activation to broadly assess how immune-related genes and regulatory pathways were perturbed by radiation.

#### The Tumor Inflammation Signature (TIS) depicts heterogeneity in prostate cancer response to HBRBT

To gain a better understanding of the functional state of the immune system in the biopsies, we first employed a modified investigational 18-gene expression profile (GEP) known as the Tumor Inflammation Signature (TIS) [15]. This signature identifies the presence of an inflamed immune infiltrate in tissues associated with activation of the IFNγ pathway, antigen presentation and T cell activation – and classifies tissues into three broad categories: high, intermediate, or low [16]. The TIS is also associated with an inflamed but immunosuppressed phenotype in many different cancer types that is also correlated with good responses to checkpoint blockade [16]. We used a modified TIS signature containing 16 of the 18 genes available from the full signature. We used hierarchical k-means clustering of the z-score-normalized expression data of the 16 genes in 46 biopsy samples to delineate categories based on the TIS genes. The results of this analysis, shown in **Figure 2A and B**, revealed a smooth distribution of the 46 pre-HDRBT samples over the three clustered categories (19 low, 14 intermediate, and 13 high). These three immune gene expression profile (GEP) categories were driven by discrete clusters of genes as indicated by the dotted boxes in **Figure 2A**. These gene clusters included increased expression of IFN-γ response genes (e.g. *CXCL9, PDL1, STAT1* and *PSMB10* bottom box), as well as CD8^+^ T cell infiltration, co-stimulation and chronic activation genes (*CD8A, TIGIT, Lag3, CD27, CCL5, CXCR6* upper box). The TIS-high category samples had increased expression of IFN-γ response and chronic T cell activation genes; in contrast, the TIS-intermediate samples had increased expression of the IFN-γ response genes only. The TIS-low samples had no evidence of an IFN-γ response or T cell activation **(Figure 2A)**. Importantly, this heatmap depicts the pre-HDRBT samples and their change in TIS category post-HDRBT, shown as white circles (low TIS), orange circles (intermediate TIS) and red circles (high TIS) **(Figure 2A)**. A more extensively annotated heatmap, including clinical characteristics, is also provided in **Supplementary Figure S5**. Prior to HDRBT, only 34.8% of the tissues were classified as either high or intermediate TIS - with 65.2% (15/23) of the biopsies being classified as low TIS. Following HDRBT, we observed a statistically significant (Chi-squared test; *p* = 0.008) increase in the proportion of tissues harboring a high or intermediate class TIS signature (82.6%; 19/23 tissues) (**Figure 2C**). Following radiation, the overall mean TIS expression significantly increased post-HDRBT, with only 4/23 (17.4%) patients exhibiting a low TIS score after HDRBT (**Figure 2D**). We also confirmed that the HDRBT-induced PCa TIS increase was patient-specific and not stochastic (**Supplementary Figure S6**). We then focused our analysis on the pre-HDRBT low TIS samples and found the vast majority (80%; 12/15) were converted to either an intermediate TIS (46.7%) or high TIS (33.3%). The remaining three patients did not respond to the radiation in terms of TIS (RA014, RA025 and RB050), with no clear underlying clinical (e.g. Gleason Grade) or experimental cause (**Supplementary Figure S6**). A bioinformatics analysis suggested that latent immune activation in baseline tissue (e.g IFNγ and TNFβ pathways) was associated with a good TIS response to HDRBT (**Supplementary Figure S7**).

**Figure 2:**
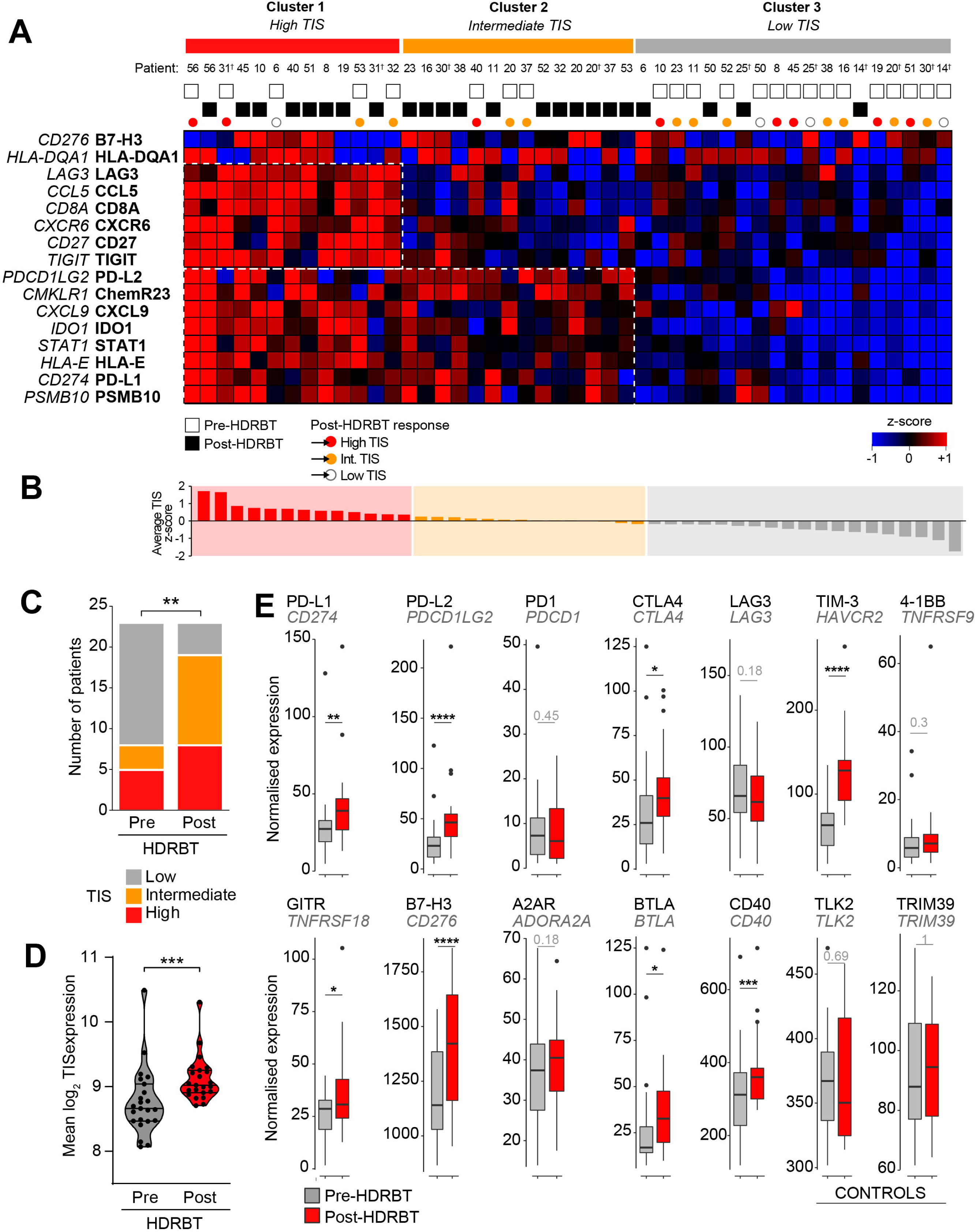
**High dose rate brachytherapy induced a 16-gene tumor inflammatory signature (TIS) in localized PCa**. Shown is (A) Heatmap of normalized expression levels of 16 genes in tumor inflammatory signature (TIS) and categorisation by k-means clustering into three groups: (i) Cluster 1, high TIS, (ii) Cluster 2, intermediate TIS, and (iii) Cluster 3, Low TIS. White and black boxes indicate either pre-HDRBT or post-HDRBT tissue, respectively. Colored circles indicate pre-HDRBT samples and their TIS category change post-HDRBT. (B) Averaged and ranked z-scores for the 16 genes in TIS indicated three categories. (C) Proportion of pre- or post-HDRBT tissues in each of the three TIS categories identified in (A), the P-value was calculated from chi-squared test. (D) Violin plot of mean TIS expression levels in patient-matched pre- or post-HDRBT-treated PCa tissue. Wilcoxon matched pair test. *p<0.05, ** p<0.01, ***, p<0.001, ****p<0.0001. (E) Box- and-whisker plots of expression levels of immune checkpoint molecules in pre- and post-HDRBT tissues from all patients in cohort. *TLK2* and *TRIM39* are provided as invariant controls. Significance was assessed using a Wilcoxon matched pair test. *p<0.05, ** p<0.01, ***, p<0.001, ****p<0.0001. † represents RadBank-V1.

Immune checkpoint (IC) molecules were significantly changed (Paired Wilcoxon test; *p*<0.001, **Figure 2E**) in response to HDRBT, including genes encoding PDL2, TIM-3, B7-H3, PDL1, CTLA4, GITR, BTLA and CD40. HDRBT-unresponsive IC molecules included PD-1, LAG3, 4-1BB and A2AR. TGFβ (in the form of its mRNA transcript *TGFB1)* is also strongly and significantly upregulated following radiation (**Supplementary Figure S6B**).

#### Immunotranscriptomic profiling the response of PCa to HDRBT

To more broadly describe immune gene expression changes induced by HDRBT, we interrogated all 770 genes evaluated by the Nanostring nCounter PanCancer Immune Profiling platform. Using a two-sample t-test, we identified 59 highly significant (false discovery rate = 0) genes that were differentially expressed in response to HDRBT (**Supplementary Figure S8A**). More in-depth analysis of these candidates revealed the strong overexpression of the p53 pathway and DNA damage-related genes (e.g. *CDKN1A*/p21 and *BAX*) (**Supplementary Figure S8B**). The monocyte/macrophage markers *CD163* and *CD14* were also highly expressed genes – both were identified in our previous pilot studies [11-13]. Amongst the T cell specific markers, we identified *LILRB2, CD86* and *IL2RA* - the latter of which is typically expressed on CD4^+^FOXP3^+^ Tregs and activated CD4^+^ T cells. Gene pathway analysis of the 59 candidates using EnrichR confirmed the enrichment of genes associated with CD14^+^ monocytes and the CD33^+^ myeloid lineage (**Supplementary Figure S8C**: Human Gene Atlas database), and macrophage, inflammation and complement activation markers (**Supplementary Figure S8D**: Wiki Pathways). We identified the immune response was diverse and included the four main immune pathways assessed by the gene panel (innate, humoral, adaptive and inflammatory; **Supplementary Figure S8E**). Importantly, we verified that changes in gene transcripts encoding the mIHC markers used for Opal analysis reflect the overall density changes observed via multiplex IHC in later results – which was an important cross platform validation of our findings (**Supplementary Figure S9**).

Taken together, our gene expression data suggests that HDRBT induced inflammation (as measured by TIS) exists in the majority of samples. However, this gene expression data is derived from bulk RNA, with several samples not harboring tumor zones. This supports the concept that the overall response is at least partially driven by the surrounding stromal tissue compartment rather than explicitly by tumor cells alone. Assessing these changes at a more detailed cellular level is crucial for unravelling the role of different immune cell types and tissue areas in driving differential responses to HDRBT. Therefore, we next employed digital spatial gene and protein expression profiling to understand this response in more detail in representative patients.

### Digital spatial profiling identifies tissue-specific TIS changes driven by complex immune infiltrate

We then explored the cellular source of TIS expression, its dependency on the presence of tumor, and the cause of the different two TIS expression patterns (i.e. intermediate and high). To do this, we selected two patients that exhibited either an intermediate (RB023) or high (RB019) TIS response to HDRBT (**Figure 3A**) and used Nanostring digital spatial profiling to determine TIS expression. IHC staining was performed with DAPI, CD3, CD68 and Pan-cytokeratin targets to define cell distribution (**Supplementary Figure S10A**) prior to regions of interest (ROIs) selection. Twelve ROIs were selected per patient sample (24 total) to include either tumor or non-tumor zones (**Supplementary Figure S10B**), and to contain a variety of T cell and MΦ infiltration (**Supplementary Figure S11**) (marked by CD3^+^ or CD68^+^, respectively). Using matched ROIs in adjacent serial sections, we performed either RNA or protein IO (immuno-oncology) analyses using 78-plex RNA or 55-plex protein DSP IO panels, respectively. We first evaluated the relative expression of TIS in each patient ROI (**Figure 3B**). K-means hierarchical clustering identified two groups of ROIs (high or low) on the basis of total TIS expression in both patients. This analysis did not completely segregate non-tumor and tumor-bearing ROIs. We also confirmed that minor differences in surface area of the ROIs did not affect TIS expression (**Supplementary Figure S10C**). To further explore the nature of TIS expression, we profiled the 24 ROIs for mean TIS expression and identified the highest and lowest-expressing tissue sites. This analysis, shown in **Figure 3C and D**, revealed that the top TIS-expressing sites in both patients were in non-tumor zones. This analysis also revealed the highest TIS ROIs **(Figure 3C-D, upper panels)** also contained a large gross immune infiltrate characterized by both CD3^+^ T cells and CD68^+^ MΦ. The lowest TIS ROIs were more variable between the patients. RB019, an overall high-TIS class tissue, exhibited low TIS expression in tumor sites irrespective of immune infiltrate levels (**Figure 3C; lower panel**). Conversely, low TIS ROIs in RB023 (intermediate TIS responder) were marked by a combination of tumor and non-tumor sites, yet largely absent T cell and MΦ infiltrate (**Figure 3D; lower panel**). To more robustly define this difference, we directly compared the mean ROI TIS levels between non-tumor and tumor sites for both patients (**Figure 3E**). This analysis revealed that patient RB019 exhibited significantly higher TIS expression in non-tumor zones, whilst RB023 indicates a similar but not statistically significant difference. This analysis also confirmed that post-HDRBT tissue in RB019 has a higher overall TIS activation as a whole when compared to RB023 (*p*<0.0001) - consistent with the analysis in **Figure 2**.

**Figure 3:**
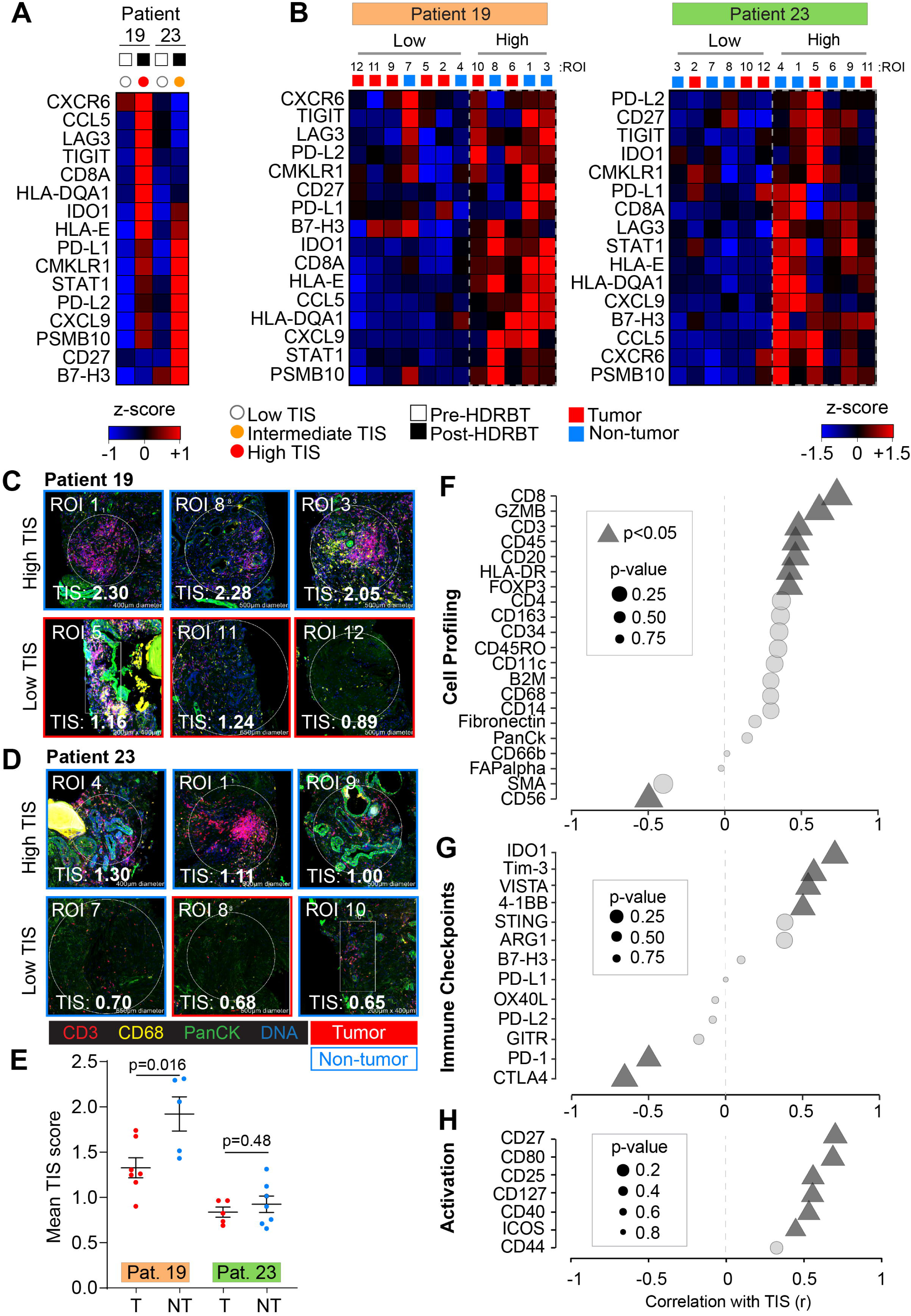
**Digital spatial gene and protein expression profiling reveals cellular and immune checkpoint drivers of high and intermediate TIS**. Heatmaps of normalized Nanostring TIS gene z-scores in two patients using **(A)** bulk tissue from pre- and post-HDRBT tissues, or **(B)** post-HDRBT ROIs (n=24) assessed using digital spatial profiling human IO RNA panel (DSP RNA). Tissue sites were determined using tumor histopathology (tumor and non-tumor) and immune cell IHC staining for T cells (CD3) and MΦ (CD68) (12 ROIs per patient). High magnification examples of highest and lowest TIS-expressing ROIs in post-HDRBT tissue from patient **(C)** RB019 and **(D)** RB023. **(E)** Mean Nanostring DSP-derived TIS scores by tumor category in two patient samples (tumor zones: n=7/12(RB019) and n=5/12(RB023)). Pearson correlation of DSP IO proteins in **(F)** cell profiling, **(G)** immune checkpoint, or **(H)** activation protein classes with DSP RNA TIS score.

To understand which immune proteins are linked with TIS expression, we correlated the DSP protein data (55 proteins) with RNA TIS scores for matched ROIs in the full set of 24 ROIs. The protein results were also subdivided into cell profiling **(Figure 3F)**, immune checkpoints **(Figure 3G)**, immune activation **(Figure 3H)**, and pan-tumor markers **(Supplementary Figure S10D)** categories to identify the primary molecules in each group. Using all 24 ROIs, we detected positive associations between markers of CD8^+^ T cells (CD45, CD3, CD8 and GZMB), MHC Class II (HLA-DR), Tregs (FOXP3), and B cells (CD20). Interestingly, we also identified a strong inverse correlation between CD56 protein levels (primarily a marker of NK, γδT cells and activated CD8^+^ T cells) and TIS. We next investigated if IC protein levels may be related to TIS. VISTA, IDO1, 4-1BB and Tim-3 were all positively associated with TIS **(Figure 3G)**. Conversely, CTLA4 and PD-1 were higher in areas of low TIS, which may indicate activated Tregs and corresponding immunosuppression. Importantly, many of these molecules were also shown to be upregulated overall in the tissue following HDRBT in **Figure 2E**. Finally, we interrogated associations with proteins involved in immune activation and co-stimulation **(Figure 3H)**. Similar to previous results, we identified that CD27, CD80, CD25, CD127, CD40 and ICOS were all positively correlated with TIS.

In summary, the DSP data in two cases suggests that the HDRBT-induced TIS we observed in **Figure 2A** is broadly driven by a mix of infiltrative immune cells that includes CD8^+^ T cells, Tregs, macrophages and DCs. Immune-absent surrounding tissue possessed a relatively low TIS. Moreover, TIS responses were more pronounced in the case with a high TIS category (patient RB019) in non-tumor areas. Such areas also do not seem to correlate with any other cell types aside from CD8^+^ T cells. Alternatively, intermediate TIS zones (patient RB023) appeared to be unaffected by the presence of tumor and possess a highly diverse immune context (T cells + Mφ + DC etc.). Our results also suggest that TIS levels were linked with IC expression including IDO1, VISTA and Tim3, and potentially inhibited by PD-1 and CTLA4.

### HDRBT-induced immune cell density alterations in PCa are dominated by regulatory T cells, DCs and macrophages

The next stage of the study used multiplexed immunochemistry (mIHC) and computational analysis of major immune cell types and how they are altered following HDRBT in PCa. In order to further understand the impact of radiation on the immune contexture of PCa, we first evaluated cell density (cells per mm^2^) for all subsets in all tissue regardless of the presence of tumor. The results, shown in **Figure 4A**, revealed that the overall density of immune cell subsets was not affected by radiation. The dominant cell populations present in the tissue were T cells, primarily comprised of CD4^+^ T cells and DNTs. Minor cell populations included PDL1^+^ macrophages and DCs, Tregs and CD8^+^ T cells. Statistical analysis revealed that Tregs and PDL1^-^ macrophage densities were significantly increased post-HDRBT.

**Figure 4:**
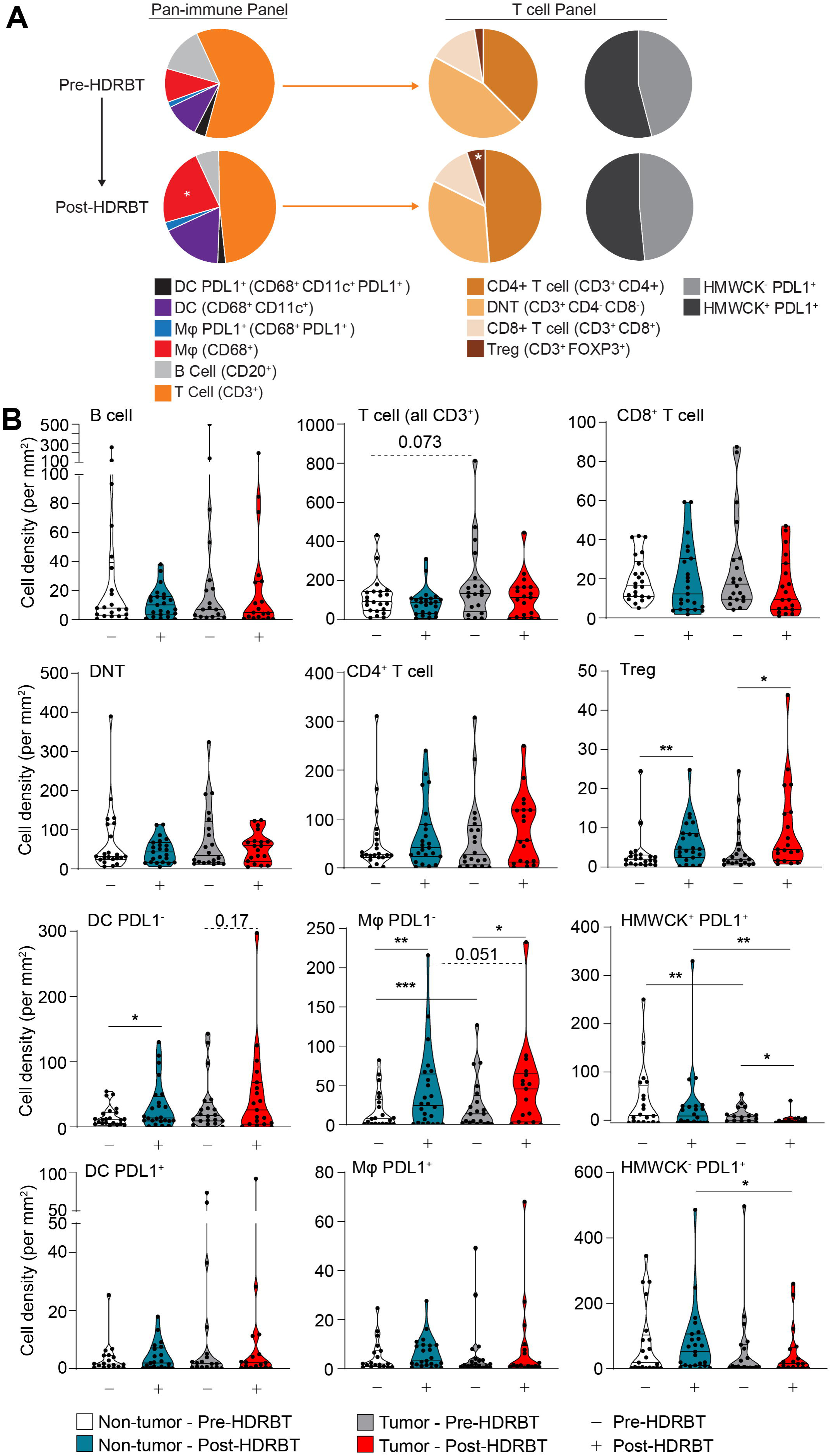
**High dose rate brachytherapy significantly increased the density of CD4**^**+**^**FOXP3**^**+**^ **T cells and antigen presenting cells**. Multiplex IHC was performed on PCa biopsies pre- and post-HDRBT (n=24 patients). Shown are (A) Cumulative barplots of mean immune cell density either pre- or post-HDRBT. P-values calculated from two-tailed student’s t-test with * indicating p< 0.05. (B) Immune subset density calculated from total number of identified immune phenotypes (see Table 2) per square millimetre pre- or post-HDRBT in each of the two designated tissue zones: (i) non-tumor or (ii) tumor-containing. Statistical significance was calculated using a non-parametric Wilcoxon signed-rank test. P-values are indicated were appropriate. *p<0.05, ** p<0.01, *** p<0.001.

As radiation is hypothesized to drive differential immune responses in prostate tumor tissue, we next assessed the impact of the presence of tumor on the immune context. The results revealed that the changes established by looking at whole tissue (i.e. Tregs and PDL1^-^ macrophages) were largely maintained in both tumor and non-tumor zones (**Figure 4B**). However, we did observe that PDL1^-^ DCs exhibited an increase in density post-HDRBT in both non-tumor (p <0.05) and tumor zones (p = 0.17). PDL1^-^ macrophages exhibited the most interesting pattern of infiltration, with a small but statistically robust density increase that was higher in tumor zones compared to non-tumor - potentially driven by the preferential phagocytotic response to tumor cell damage and apoptosis following HDRBT. PDL1^+^ APCs (Mφ and DC) were a very minor population and exhibited no change in response to HDRBT. As expected, the density of HMWCK^+^ cells was lower in tumor zones and highly sensitive to HDRBT. PDL1^+^ HMWCK^-^ cells, a mix of tumor and normal stromal cells were uniquely decreased in tumor zones. The decreased density of stromal/tumor cells that express PDL1, along with increased PDL1-expressing APCs in the tumor sites, is potentially indicative of zone-specific cell death from ionizing radiation (e.g. DNA damage-induced apoptosis), inflammation, and subsequent recruitment of macrophages to the irradiated site. The results of this analysis revealed the immune cell density in prostate tissue is heterogeneous and surprisingly resilient to HDRBT; evident in only modest changes in cell densities.

### The Tumor inflammation signature is associated with radioresponsive immune cell relationships

Localized immune network signaling occurs following key interactions between immune cells (e.g. antigen presentation between DCs and cytotoxic T cells). We hypothesized that HDRBT also alters relative density associations as well as spatial features (i.e. distance) between key immune cell subsets. Similar to the overall density of certain immune subset, this is also hypothesized to influence TIS activation levels.

#### Cell density associations in non-tumor and tumor zones are perturbed by HDRBT

We first performed an analysis of the correlations between different immune subsets in all samples and included both tumor and non-tumor tissue zones. This revealed that numerous immune cell densities were significantly correlated in our PCa cohort (**Supplementary Figures S12A and S12B**) regardless of tumor content. These subset pairs include T cells/B cells, CD4^+^ T cells/Tregs, and PDL1^-^ APCs (DC and Mφ) with CD4^+^ T cell subsets (CD4+ T cell and Treg). When cell densities were subdivided by tissue zone and irradiation status, only a proportion of these associations are retained in common (e.g. CD4+ T cell and Treg) and most were restricted to each tissue type without a clear pattern (**Supplementary Figure S12C)**. We therefore performed a differential Pearson correlation analysis to better resolve these differences. The results of this analysis (**Figure 5A**) revealed only two association gains were conserved between the two tissue zones, between PDL1^+^ DC and PDL1^+^ Mφ, and Tregs and B cells. The strongest post-radiation changes in tumor zones only included a gained association between T cells and PDL1^+^ HMWCK^+^ cells. Interestingly, this occurred in parallel with lost associations between T cells and both PDL1^+^ macrophages and B cells. Following HDRBT, both Tregs and CD4^+^ T cells were more associated with PDL1^+^ HMWCK^-^ in the tumor zone only. The interaction of PDL1^-^ DCs with their PDL1^+^ counterparts, as wells as with PDL1+ macrophages and B cells, were also preferentially lost in tumor zones. Taken together, this analysis renders a picture of radiation-induced changes in associations between T cells and APC subsets that appears to be influenced by the presence of tumor.

**Figure 5:**
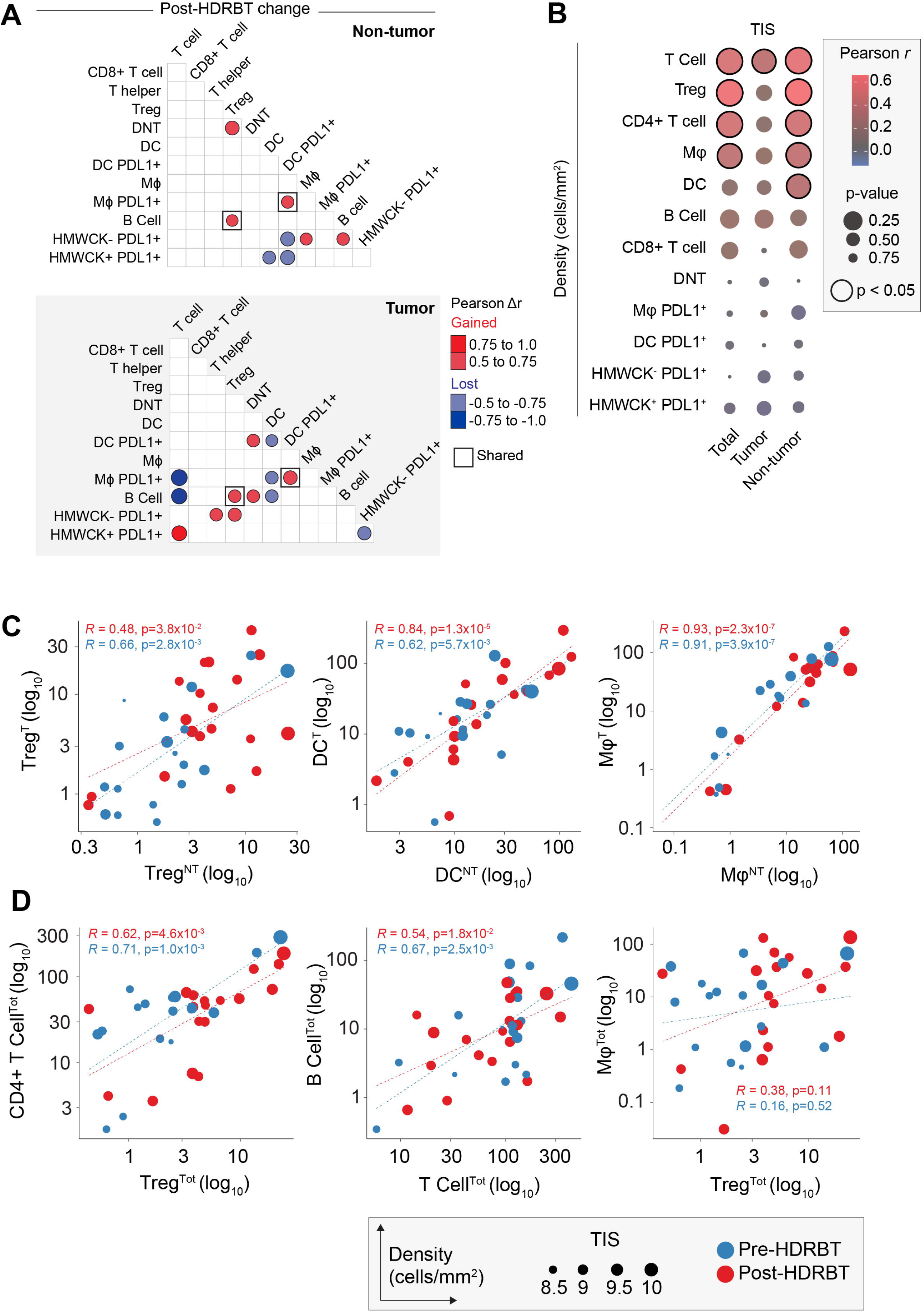
**Immune cell density relationships correlate with TIS signature**. **(A)** Computed differences in Pearson correlation (Δr) for either (i) non-tumor and (ii) tumor following HDRBT. Color indicates direction of change, i.e. red = gained association, blue = lost association. Only significant associations are shown **(B)** Bubblechart of correlation (Pearson *r*) between immune cell densities in different tissue zones (total, tumor or non-tumor) and TIS signature. Significant correlations indicated. **(C)** Scatterplots of highly significant immune cell subset correlations between tumor and non-tumor tissue zones, and **(D)** highly significant cross-subset correlations in total tissue, with relative TIS level and HDRBT radiation-status indicated. (T: tumor zone, NT: non-tumor zone, Tot: all tissue zones). Pearson correlation *r* values and corresponding *p* values indicated.

To understand if the densities of immune subsets correlate with changes in tumor inflammation (TIS), we next correlated total, tumor and non-tumor immune subset densities with the TIS signature using Pearson *r* analysis. The results (**Figure 5B and Supplementary Figure S13**) confirmed that CD3^+^ T cells, Tregs, CD4^+^ T cells and Mφ all correlate significantly with TIS. Perhaps most interesting was that these correlations were most significant in non-tumor tissue, with the exception of CD3^+^ T cells. DC’s were also observed to correlate with TIS in non-tumor zones. Overall, this suggests that immune cells in non-tumor areas are responsible for changes in TIS. To support this, we next compared the relative density of Treg, DC and Mφ between tumor and non-tumor zones (**Figure 5C**). This analysis confirmed that all were strongly correlated, suggesting other factors are likely involved. These data suggest many immune subsets are strongly correlated, and also responsive to radiation in terms of TIS. We therefore compared the most potent combinations (**Figure 5D**) and revealed that some are highly correlated (CD4^+^ T cells/Tregs and B cell/T cell) and some non-correlated (Treg/Mφ). Overall, this density analysis reinforces the notion that specific combinations of immune cell relationships are more important to the HDRBT TIS response than others.

#### Tumor-specific immune cell distance relationships are tightly linked with the TIS response to HDRBT

Another major determinant of immune signaling in tissue is the spatial relationships between key immune cell subsets. To address this, we used the spatial information within the multiplex IHC dataset to characterize the median distance between all immune cell subsets (**Supplementary Figure S14**). This resulted in a total of 72 unique interactions over the two multiplex IHC panels in each of the four tissue types according to tumor and radiation status. We first used ANOVA analysis to shortlist those spatial relationships with statistical difference, and identified seven with some degree of variation between the four groups (**Supplementary Table 3**). The most significant changes were observed between CD8^+^ T cell/CD8^+^ T cell (*p* < 0.00097), T cells/DCs (*p* < 0.0048), and Mφ/DC (*p* < 0.0072). Other significant differences were observed between DNT/Treg, B cells/DC PDL1^+^, CD4^+^ T cells/CD8^+^ T cells, and HMWCK^-^PDL1^+^/ CD8^+^ T cells. We next performed a statistical pairwise Wilcoxon test analysis to resolve specifically where these changes occurred (**Figure 6A**). We revealed that the main spatial response to radiation was a decrease in distance between T cells/DC and Mφ/DC decreases in both tumor and non-tumor zones. Conversely, the only spatial proximity which was lost was between CD8^+^ T cells as a whole. We also observed that DNT and Tregs were closer together in tumor areas.

**Table 3:** Top 20 significantly perturbed median spatial relationships in PCa tissue in response to HDRBT.

| *Target* | *Neighbour* | *ANOVA p-value* |
| --- | --- | --- |
| *CD8+ T cells* | CD8+ T cells | **0.00097** |
| *T cells* | DC | **0.0048** |
| *MΦ* | DC | **0.0072** |
| *DNT cells* | Tregs | **0.018** |
| *B cells* | DC PDL1+ | **0.04** |
| *CD4+ T cells* | CD8+ T cells | **0.04** |
| *HMWCK-PDL1+* | CD8+ T cells | **0.046** |
| *T cells* | MΦ | 0.056 |
| *DNT cells* | CD8+ T cells | 0.06 |
| *T cells* | DC PDL1+ | 0.081 |
| *HMWCK-PDL1+* | Tregs | 0.095 |
| *CD8+ T cells* | HMWCK-PDL1+ | 0.1 |
| *MΦ* | DC PDL1+ | 0.11 |
| *HMWCK+PDL1-* | CD8+ T cells | 0.11 |
| *CD4+ T cells* | CD4+ T cells | 0.14 |
| *Tregs* | CD8+ T cells | 0.16 |
| *HMWCK-PDL1+* | CD4+ T cells | 0.16 |
| *B cells* | DC | 0.17 |
| *CD4+ T cells* | HMWCK-PDL1+ | 0.18 |
| *DNT cells* | HMWCK-PDL1+ | 0.22 |

**Figure 6:**
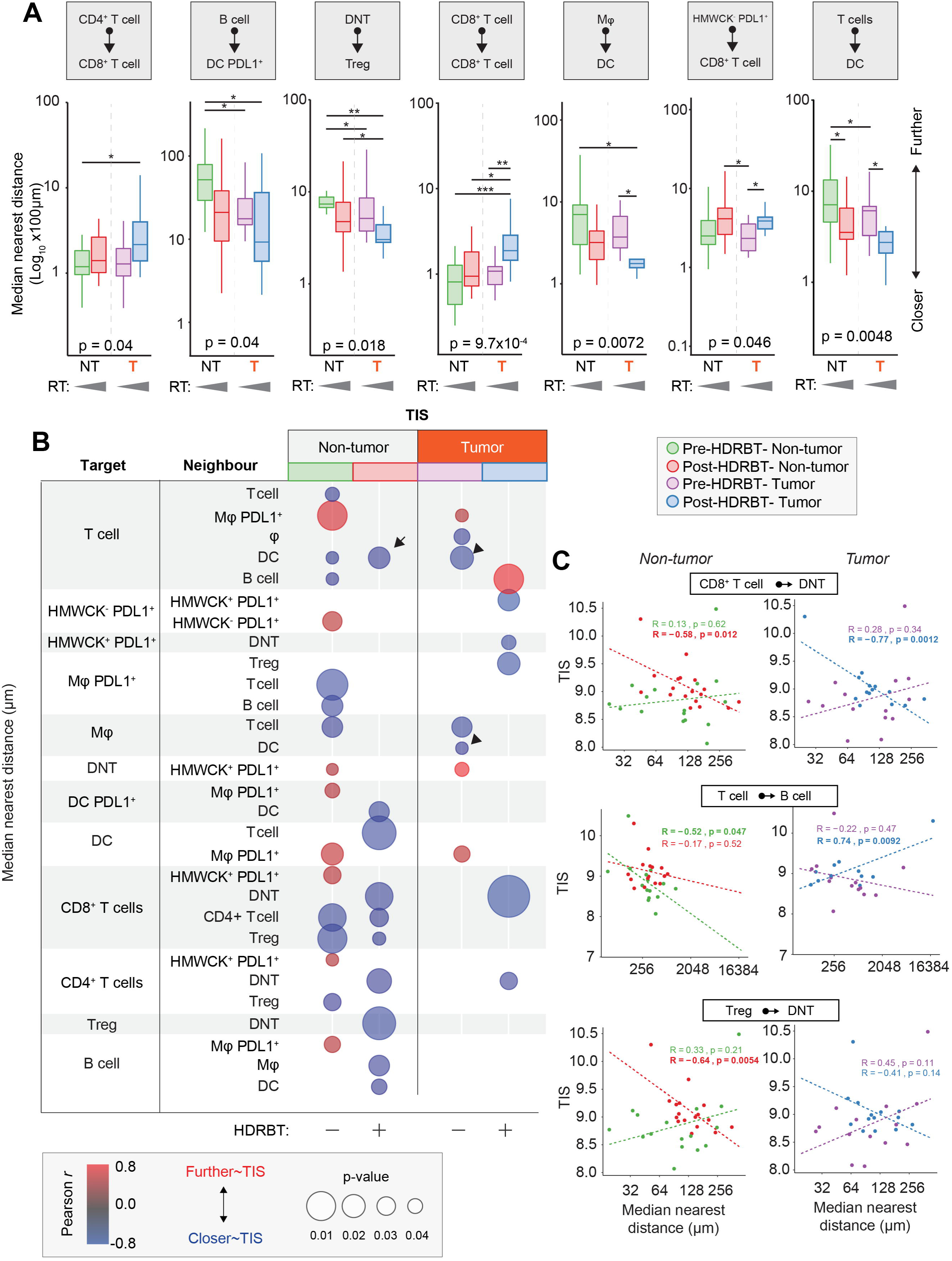
**Immune cell spatial interactions change following HDRBT and uniquely associate with TIS signature**. **(A)** Box-and-whisker plots of median cell-cell distance in seven significantly altered immune cell subset pairs in four major tissue zones: (i) Pre-HDRBT: Non-tumor zone, (ii) Post-HDRBT: Non-tumor zone, (iii) Pre-HDRBT: Tumor zone, and (iv) Post-HDRBT: Tumor zone. Significance between group is calculated using uncorrected Wilcoxon test. n=17 patients. **(B)** Bubblechart plot of significant target- and neighbour-cell median distance correlations with TIS signature in four tissue groups. Positive correlations indicate a higher median cell-cell distance association with higher TIS. Only significant (p<0.05) correlations are shown. **(C)** Examples of highly significant median distance to TIS correlations with Pearson *r* calculations for each of four categories of tissue. *p<0.05, *p<0.01, ***p<0.001.

Similar to the previous density analysis, we next compared the median distance between all 72 subsets and the normalized TIS expression level. The results, shown in **Figure 6B**, showed that 29/72 interactions were influenced TIS. There was a significant difference according to tumor status, with 67% and 33% being associated with TIS in non-tumor and tumor zones, respectively. This corroborates our previous density analysis which suggested that the non-tumor areas are most important for TIS activation. Also, the bulk of these correlations (70%) were negative, largely in post-HDRBT tissue (100% of non-tumor, 83% of tumor zones). Some of the most significant examples of these changes are provided in **Figure 6C**. Overall, this analysis suggests that close cell-cell interactions are key for increased TIS activation following HDRBT. However, we did observe one post-HDRBT increase at a distance that was specific to tumor zones, between T cells and B cells.

## Discussion

Clinical outcomes for patients with localized PCa are linked to features such as the serum prostate-specific antigen level, tumor stage and grade, along with treatment factors. Patients with more aggressive clinical features however have higher rates of recurrence with all treatment options [1, 4], including with HDRBT, and these recurrences may be either local or metastatic [17] and significantly impact survival [18].

Key to improving outcomes from HDRBT is the detailed understanding of cellular and molecular responses to the radiation, with a view to design of rational combination approaches. Here we have specifically explored HDRBT-induced immune responses in PCa at the level of gene expression signatures and infiltrative immune cells, and also asked whether these changes were driven by tumor cells or surrounding tissue. This is the first study to analyze immune responses to HDRBT in localized human PCa. The immune context of localized PCa has thus far only been explored in cells isolated from radical prostatectomy samples [6, 19-23]. Prior studies showed PCa infiltrating lymphocytes (PILs) included CD8^+^ effector memory T cells [21] which were oligoclonal and PD-1^+^ [22], accompanied by increased regulatory CD4^+^ and CD8^+^ T cells with suppressor function [19, 20]. In addition, IHC showed PCa lesions were surrounded by lymphocyte clusters enriched for FOXP3^+^, PD-1^+^, and PDL1^+^ cells [6]. Thus, the emerging picture for PCa is one of immune exclusion and immune suppression.

Our results revealed that PCa tissues have immune activation states in three distinct TIS levels with two-thirds of pre-HDRBT tissues having a low TIS - indicating an inactive immune microenvironment. This distribution was reversed following radiation, with over 80% of tissues possessing a high or intermediate TIS after HDRBT. Using a combination of multiplex IHC and digital spatial gene and protein profiling (DSP), we showed this TIS change is intimately linked with several immune cell densities, specifically those of active T cell subtypes and APCs - supporting the notion that the two processes are linked. The DSP analysis also revealed that the strongest TIS responses are driven primarily by non-tumor zones, and was associated primarily with CD8^+^ T cells. Conversely, intermediate TIS is agnostic to tumor and is linked with a much more diverse group of T cell and APC cell subtypes. The presence of numerous therapeutically significant IC molecules in conjunction with TIS in two selected cases, including VISTA, IDO1, 4-1BB and Tim3, raises possibilities for combination treatment with HDRBT. Both VISTA and IDO1 were associated with tumor resistance in ICB-treated prostate cancer [24, 25]. Whether these IC molecule changes are drivers or responders to HDRBT-induced responses remain to be seen but warrants further investigation in additional patients and radiation states. Overall, these results reveal that immune signaling gene expression is profoundly influenced by HDRBT and is not confined to tumor areas. A caveat of our study is that the biopsy procedure, in conjunction with HDRBT, could contribute to inflammatory and wound healing responses in the tissue.

To further understand the potential cellular drivers of this response, we used multiplexed IHC to characterize the immune cell environment before and after HDRBT. This revealed that Tregs, Mφ and DCs were the predominant cellular density change in response to HDRBT. TGFβ, which was highly expressed post-radiation is also an immunosuppressive cytokine in prostate cancer and linked with the presence of Tregs and Mφ [26]. This occurred in the context of the majority of other immune cell types which did not alter in density, yet likely repopulated the TME to a similar level following the ablative dose of radiation. Interestingly, DNTs represented a relatively large fraction of T cells in both pre- and post-HDRBT conditions (∼35% total T cells). Whilst interesting, our analysis could not further resolve this population due to the absence of appropriate markers for putative sub-types of DNTs (e.g. γδT cells, MAIT cells, NKT cells). Despite relatively few cells significantly changing in overall density, we observed numerous cell association changes following radiation, predominately between PDL1^+^ APCs and T cell subsets. For Tregs and DCs, this effect was largely observed to occur in all areas of PCa tissue. However, the density of PDL1^-^ macrophages is positively influenced by the presence of tumor– potentially indicating the presence of M2 polarized tumor-associated macrophages (TAMs). This observation was supported by the Nanostring gene expression profiling which revealed markers of M2 TAMs [27], notably CD14, CD163, HLA-DRA and CD68, were enriched post-HDRBT. This observation was also noted in our existing proteomic analyses of post-HDRBT PCa tissue [13]. The presence of large numbers of T cell subsets after this substantial dose of radiation may indicate trafficking of new T cells.

Immune checkpoint molecule expression in cancer tissue is of paramount importance to the efficacy of immune checkpoint blockade drugs [28]. We identified a large number of changes in many IC molecules, and all increased following HDRBT. Despite not being certain of the cellular source of these molecules, increased PDL1 and PDL2 expression is consistent with the observed increases in M2 macrophages and DCs, however, our multiplex IHC analysis of PDL1 expression on these cells does not support this conclusion. We instead observed *decreases* in PDL1^+^-expressing HMWCK^+^ (basal glands) and HMWCK^-^ (tumor, epithelial, fibroblast) cells after HDRBT. This suggests that the source of increase in PDL1 expression is likely to be from cells that were not assessed by mIHC, either by differences in antibody clone specificity or simply that its expression is tightly controlled at the translational level. CTLA4 and TIM3 are both highly expressed on Tregs [29, 30], and high expression of B7-H3 and CD40 are both observed on licensed APCs. Further work will be required to understand the source of these IC molecule changes and to understand how these changes are relevant to the immune response post-HDRBT.

A striking observation made through this study related to the changes in the magnitude and diversity of spatial relationships between different cells in both tumor and non-tumor regions in response to HDRBT. Whilst many associations were acutely affected by radiation (e.g. increased proximity of T cell and DCs), further studies exploring T cell function are needed to reveal what this means to the immune response in PCa. These spatial relationship changes could dramatically affect intrinsic immune signaling pathways, we observed that many tissue-specific spatial interactions were strongly linked with TIS signatures (e.g. loss of T cell – B cell proximity in tumor zones). Our DSP data suggests that localized factors either control or are responsive to the TIS response. Importantly, patients appear to exhibit very different responses to radiation. Further work will be required to unravel the clinical importance of these relationship, if they are of predictive value and relevance to therapies.

Radiation therapy has the potential to change the solid tumor microenvironment to make this amenable to trafficking and the persistence of T cells. We demonstrated that HDRBT-induced changes in the immune cell density of PCa are restricted to macrophages, DCs, and Tregs. This suggests the observed changes in TIS levels post-HDRBT are the consequence of altered immune cell network signaling, and was correlated with changes in immune subset spatial relationships. This suggests that a significant number of PCa patients have a pre-existing immune response, which is held in check by peripheral tolerance mechanisms. Likely candidates in PCa include high levels of TGF-β and increased Tregs and immune-suppressive macrophages.

In terms of the clinical relevance of our findings, existing GEP studies have revealed that Th1-mediated adaptive immunity and inflammatory GEPs are positively associated with good outcome in patients with localized PCa [31]. Additional work has also focused on generating gene expression profiles associated with clinical outcomes, rather than predicting immune responses, which has ramifications for immune-targeted therapies [32].

## Conclusions

Taken together, our findings suggest that strategic therapeutic targeting of immunosuppressive mechanisms (e.g. TGF-β), plus immune checkpoint inhibitors with HDRBT, could help drive a systemic response to PCa. We envisage that this could be used to improve cancer control in those with high-risk disease where outcomes are currently poor, or to enable treatment de-intensification to improve the side-effect profile in those with less aggressive disease treated with RT. Larger numbers of patients with assessment of long-term clinical outcomes will be required to fully understand the full potential of immunotherapeutic approaches in PCa radiotherapy.

## Data Availability

The datasets used and/or analysed during the current study are available from the corresponding author on reasonable request

## List of Abbreviations

PCa: Prostate cancer
HDRBT: High dose-rate brachytherapy
TIS: Tumor inflammation signature
EBRT: External beam radiation therapy
RT: Radiation therapy
IHC: Immunohistochemistry
mIHC: Multiplexed immunohistochemistry
IC: Immune checkpoint
ADT: Androgen deprivation therapy
PMCC: Peter MacCallum Cancer Centre
MRI: Magnetic resonance imaging
PET: Positron emission tomography
ISUP: International Society of Urological Pathology
FFPE: Formalin-fixed paraffin-embedded
GG: Gleason grade
HIER: Heat-induced epitope retrieval
DSP: Digital spatial profiling
ROI: Region of interest
RB: RadBank
GEP: Gene expression profile
IO: Immuno Oncology
APC: Antigen presenting cell

## Acknowledgements

The authors would like to thank the Molecular Genomics Facility and the Centre for Advanced Histology and Microscopy at the Peter MacCallum Cancer Centre for their assistance with performing experiments.

## Figure Legends

**Supplementary Figure S1:** Multiplex IHC experimental overview and example of pipeline for quantification of immune subsets in human prostate cancer tissue biopsies. (A) Workflow of experimental multiplex IHC study. (B) H&E stain and corresponding Opal™ tissue classification of non-tumor (blue) and tumor (red). (C) Raw acquired images from either pan-immune (left) or T cell panels (right). Example high magnification (D) non-tumor zone and (E) tumor zone. (F) Deconvoluted and processed images prior to quantification of cell types. (G) Spatial plot of identified immune subtypes indicating assignment of different cell types on the tissue and their spatial relationship.

**Supplementary Figure S2:** The effect of tumor and non-tumor amount on PCa immune context. Using the segmentation data from Figure S1B, each prostate biopsy was depicted by (A) Total area of pathologically defined tumor and non-tumor (mm^2^). (B) Correlation between tumor proportion and total tissue proportion. (C) Correlation between total tissue area and total immune cell density evaluated by multiplex IHC. Corresponding r values are shown with p-values evaluated using Pearsons correlation (95% confidence interval).

**Supplementary Figure S3:** Pearson’s correlation scatterplots of CD3^+^ cell densities in assessed tissues evaluated using either Pan-immune or T cell panels. Data was divided by either (A) non-tumor or (B) tumor zones. Dots are coloured by either pre- or post-HDRBT state with corresponding Pearson’s r values and p-values shown (95% CI).

**Supplementary Figure S4:** (A) TIS score and (B) immune cell densities according to ADT-treatment status in pre-HDRBT biopsies. Paired t-tests used for statistical analysis with p-values indicated.

**Supplementary Figure S5: High dose rate brachytherapy induced a 16-gene tumor inflammatory signature (TIS) in localized PCa**. Figure 2A with inclusion of clinical characteristics. Shown is (A) Heatmap of normalized expression levels of 16 genes in tumor inflammatory signature (TIS) and categorisation by k-means clustering into three groups: (i) Cluster 1, high TIS, (ii) Cluster 2, intermediate TIS, and (iii) Cluster 3, Low TIS. Grey and black boxes indicate either pre-HDRBT or post-HDRBT tissue, respectively. Colored circles within grey boxes (pre-HDRBT) indicate post-HDRBT TIS category change. Other clinical parameters are shown above (i.e. Gleason Grade Group, T-stage, relapse status, and ADT pre-treatment).

**Supplementary Figure S6:** (A) Patient line-matched mean TIS expression levels in pre- or post-HDRBT-treated PCa tissue. Red lines indicate increase, blue lines indicate decrease. Significance assessed using Wilcoxon matched pair test. *p<0.05, ** p<0.01, ***, p<0.001, ****p<0.0001. (B) *TGFB1* mRNA levels pre- and post-HDRBT in 24 patients.

**Supplementary Figure S7: A pre-existing immune response is associated with a tumor inflammatory signature response to HDRBT**. Nanostring immune GEP data was examined on paired pre- and post-HDRBT biopsies. Good responders were classified as those that initially had a low tumor inflammation signature (TIS) pre-HDRBT (15 patients; 65% total patients), but were subsequently converted to an intermediate or high TIS post-HDRBT (n=12/15; 80% of low TIS). Poor responders were classified as those with no post-HDRBT change (n=3/15; 20% of low TIS). (A) Z-score gene expression heatmap of significant changes calculated using a t-test. (B) Bubblechart of gene set enrichment analysis using curated Hallmark datasets and (C) example summary plots from selected immune signalling pathways (bold). (D) mRNA expression levels of IFNγ, IFNα, and TNFβ in samples from two low TIS categories (R: responder and NR: non-responder), and intermediate and high TIS categories.

**Supplementary Figure S8: Nanostring immune profiling of post-HDRBT response in prostate tissue from 23 patients**. (A) Volcano plot of post-HDRBT changes in 770 changes in Nanostring panel. FDR = false discovery rate. Highly significant candidates (FDR=0) indicated in dotted line box. (B) Magnification of highly (n=57) significant candidates and colour coding according to assigned function according to Nanostring designations or subsequent gene ontology analyses. Gene ontology analysis of candidates (n=59) using (C) Human Gene Atlas or (D) WikiPathways. (E) Volcano plots of discrete immune gene changes sorted by four functional immune classes defined in the nCounter Nanostring Pan Cancer Immune Panel.

**Supplementary Figure S9:** (B) Box and whisker plots of transcript expression levels of the 8 IHC markers (*CD3E, CD4, CD8A, CD68, ITGAX, MS4A1, FOXP3* and *PDL1*) either pre- or post-HDRBT that were also assessed in multiplex IHC analysis. Differences were assessed using a Wilcoxon matched pair test. *p<0.05, ** p<0.01, ***, p<0.001, ****p<0.0001.

**Supplementary Figure S10: Digital spatial profiling supplementary information 1**. (A) 4-color (CD3, CD68, DAPI and PanCK) stains of RNA and protein slides with selected ROIS indicated. Scale bar equivalent to 2mm. (B) Flowchart of ROI selection in post-HDRBT tissue in two patients (RB019 and RB023). (C) DSP protein correlation of pan-tumor class (c.f. Figure 4F,G,H)

**Supplementary Figure S11: Digital spatial profiling supplementary information 2**. IHC of 24 ROIs selected for RNA and protein DSP IO panels. Panels are organised by TIS category determined previously (c.f. Figure 4B).

**Supplementary Figure S12:** Correlation dot plots of normalized TIS scores and immune cell population densities for all 46 tissues in cohort. Dots are coloured according to Pearson correlation coefficient value.

**Supplementary Figure S13:** (A) Pearson correlation between immune cell subsets in all biopsies (n=24) using total tissue information (tumor + non-tumor). Squares indicate important associations investigated in B. (B) Representative scatterplot correlations between immune cell subsets. 95% confidence interval from Pearson correlation. (C) Bubblechart of pearson correlations when sub-categoriesed by HDRBT status and tissue zone. Only significant (p>0.05) correlations are shown with squares indicating shared correlations between tissue zones.

**Supplementary Figure S14:** Graphical illustration of median cell distance calculation used for Figure 7.

